# Wastewater Based Epidemiology as a surveillance tool during the current COVID-19 pandemic on a college campus (East Carolina University) and its accuracy in predicting SARS-CoV-2 outbreaks in dormitories

**DOI:** 10.1101/2023.08.04.23293359

**Authors:** Avian White, Guy Iverson, LaNika Wright, John T. Fallon, Kimberly P. Briley, Changhong Yin, Weihua Huang, Charles Humphrey

## Abstract

The COVID-19 outbreak led governmental officials to close many businesses and schools, including colleges and universities. Thus, the ability to resume normal campus operation required adoption of safety measures to monitor and respond to COVID-19. The objective of this study was to determine the efficacy of wastewater-based epidemiology as a surveillance method in monitoring COVID-19 on a college campus. The use of wastewater monitoring as part of a surveillance program to control COVID-19 outbreaks at East Carolina University was evaluated. During the Spring and Fall 2021 semesters, wastewater samples (N= 830) were collected every Monday, Wednesday, and Friday from the sewer pipes exiting the dormitories on campus. Samples were analyzed for SARS-CoV-2 and viral quantification was determined using qRT-PCR. During the Spring 2021 semester, there was a significant difference in SARS-CoV-2 virus copies in wastewater when comparing dorms with the highest number student cases of COVID-19 and those with the lowest number of student cases, (*p*= 0.002). Additionally, during the Fall 2021 semester it was observed that when weekly virus concentrations exceeded 20 copies per ml, there were new confirmed COVID-19 cases 85% of the time during the following week. Increases in wastewater viral concentration spurred COVID-19 swab testing for students residing in dormitories, aiding university officials in effectively applying COVID testing policies. This study showed wastewater-based epidemiology can be a cost-effective surveillance tool to guide other surveilling methods (e.g., contact tracing, nasal/salvia testing, etc.) to identify and isolate afflicted individuals to reduce the spread of pathogens and potential outbreaks within a community.

## Introduction

The continuous systematic collection of data coupled with its analysis and interpretation describes the basic tenants of what is considered public health surveillance.^1^ Equally important is the timely dissemination of this information to necessary officials, enabling them to make appropriate decisions in response to a problem.^2^ Thus, public health surveillance systems are theoretically constructed to provide data to assist in interventions (e.g., inhibit spread of disease).^3^ Indicator-based surveillance systems often includes some type of regular data collection and weekly alert threshold monitoring.^4^ Hence, this system may be effective in monitoring weekly bacterial or viral concentrations of specified diseases.

First used as an indicator tool of community drug use, testing of untreated wastewater has progressed to measuring infectious pathogens.^5–7^ The availability of sample procurement; there are typically multiple wastewater sampling locations (i.e., manholes) and feasibility of sampling tools (i.e., autosamplers), may enhance the ability to use wastewater as a form of surveillance. Wastewater-based epidemiology (WBE) is based on the rationale that fecal and urinary biomarkers may be used to give time sensitive information on population health.^9^ Thus, it is important to determine disease pathogens that may be stable in wastewater or detectable in sewage. Hence, with the first outbreak of severe acute respiratory syndrome coronavirus (SARS-CoV) in 2003, researchers studied and identified the detectability of the pathogen in wastewater of infected hospital patients.^10^ The ability to detect SARS-CoV in wastewater has led researchers to reasonably conclude that wastewater may be used to detect to viral SARS-CoV-2 resulting from the current SARS COVID -19 outbreak.

The University of Arizona used wastewater-based epidemiology as part of a re-entry strategy for students returning to campus. Researchers there monitored wastewater in a student dormitory, where upon detection of SARS-CoV-2 resulted in clinical testing of students living in the dorm. Officials found WBE important in containing COVID-19.^11^ Similarly, an interdisciplinary team at Norwich University set out to determine if WBE could be used to assist its school in COVID-19 surveillance and detection .^12^ More universities have begun to incorporate the use of WBE as a part of their COVID-19 mitigation strategies. Schools like UNC Charlotte have sought to use the results obtained from their wastewater testing as part of their approach to stemming cases and found it to have been successful at early detection.^13^ Thus, East Carolina University implemented a WBE framework to identify dormitories with elevated SARS-CoV-2 concentrations to guide public health interventions.

The aim of this research was to evaluate the use of wastewater-based epidemiology as a surveillance tool during the COVID-19 pandemic by comparing SARS-CoV-2 concentration in wastewater to the number of confirmed cases of COVID-19 in dormitories on the campus of East Carolina University. Specifically, it was hypothesized that dorms with higher numbers of confirmed student COVID-19 cases would have statistically higher concentrations of SARS-CoV-2 in their wastewater. It was also hypothesized that sampling and analyses for SARS-CoV-2 in dorm wastewater would be effective in identifying new cases of COVID-19 in dormitories, thus potentially helping to prevent possible spread and future outbreaks of the disease.

## Methods

### Sample Procurement

Wastewater from dormitories on ECU’s campus was collected and analyzed during the 2021 Spring (9 dormitories) and Fall semesters (16 dormitories), respectively. The university utilized fewer dormitories during the Spring semester, resulting in a difference between sampled dorms between the semesters. This was attributed to classes being held predominantly online during the Spring semester and the university only allowing for single occupancy in dormitories. Thus, fewer students stayed on campus. Wastewater samples were collected Monday, Wednesday, and Friday excluding holidays and university approved days off (n= 830). Wastewater samples from each dormitory on campus were collected using Hach AS950 portable samplers with vacuum tubes extending down a manhole and into the main sewer pipe exiting each dormitory (Figure 1). Each Hach AS950 used a peristaltic pump to pull wastewater through the tubing from the sewer pipe into a 7.6-L capacity sample bottle. Sample bottles were encased in ice within the sampler housing to ensure stability of samples at 4°C. For this study, the pumps were programmed to collect 21 mL of raw wastewater every 15 minutes, equating to 84 mL per hour, which allowed for collection of a daily composite wastewater sample. On sample collection days, composite samples were retrieved from autosamplers and stored on ice until they were delivered to the Water Research Laboratory at East Carolina University.

**Figure 1.**
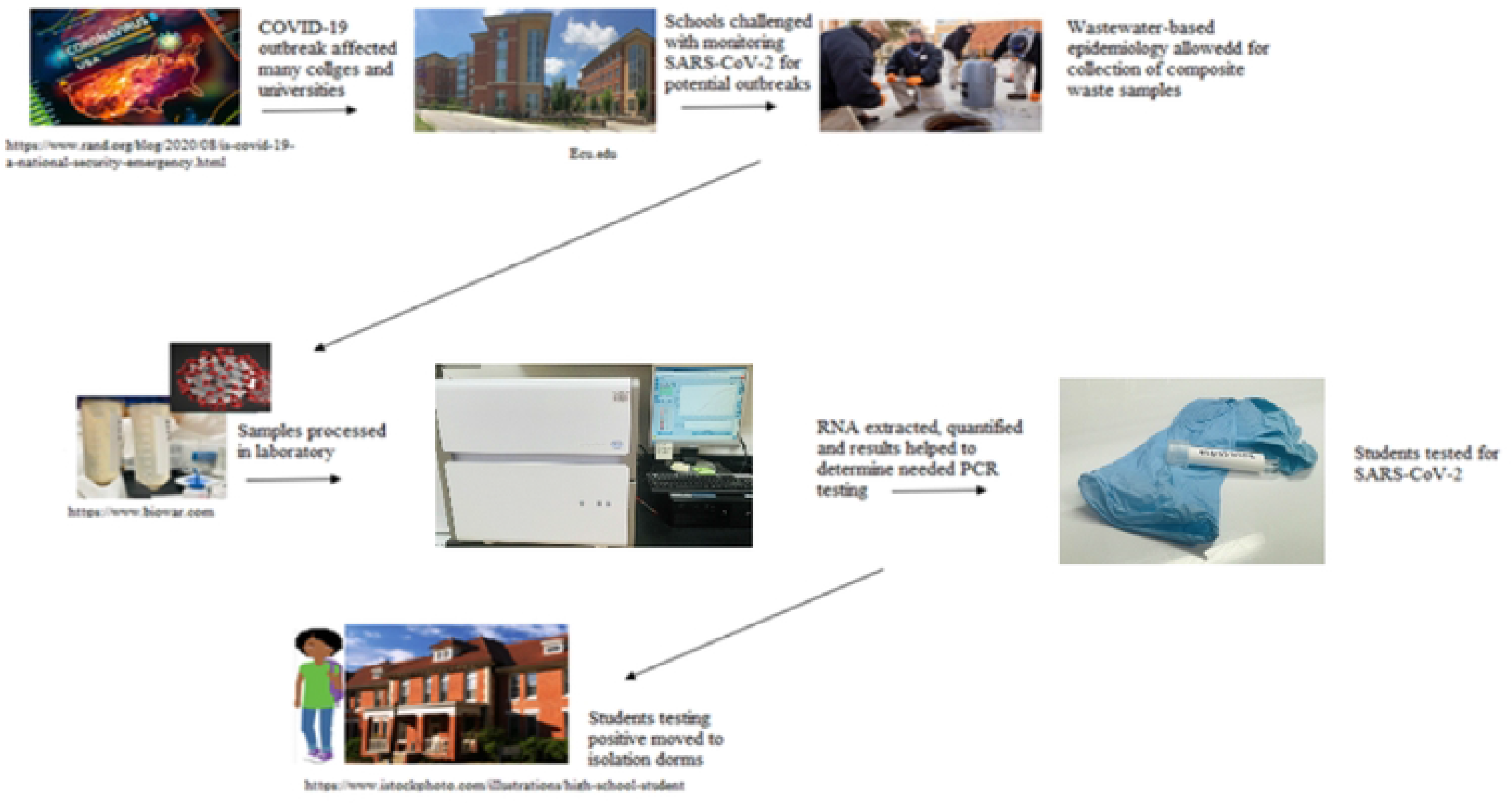
Flow Diagram of Wastewater-Based Epidemiology on Campus of East Carolina University.

### Sample Processing

Approximately 95 mL of composite sample was aliquoted into 2 labeled, 50 mL conical tubes corresponding with the respective dormitory. Samples were heat pasteurized via placement into a 75°C water bath for 45 minutes (Pecson et al., 2021; Kitajima et al., 2020; Ahmed et al., 2020). Next, samples were removed from water baths and placed into -80°C freezer to cool samples to between 2° - 8°C. Temperatures were verified using a Traceable Precision Thermometer (Fischer Scientific Cat# 150790712). Cooled samples were centrifuged at 4100 RPM for 30 minutes at 4°C, to remove large particles. The supernatant was decanted into new 50 mL ultra-centrifugation conical tubes containing a mixture of 3.5 ± 0.1 g Polyethylene Glycol 8000 (PEG) and 0.788 ± 0.01 g NaCl and mixed until the PEG/NaCl mixture was dissolved. The resultant solution was centrifuged at 12,000 × g at 4° C for 30 minutes, ensuring the formation of the viral pellet. The supernatant was carefully decanted from the conical tube so as not to disturb the viral pellet. The pellet was resuspended using 1 mL of TRIzol and transferred to the second sample tube containing a viral pellet (IDEXX, Westbrook, Maine). The resultant 1 mL solution was transferred into a labeled 2 mL microcentrifuge tube and placed on ice. Samples were transferred to a Pathology laboratory at Brody School of Medicine, East Carolina University for RT-qPCR analysis.

### Quantifying SARS-CoV-2 Concentration using RT-qPCR

RNA was extracted from PEG8000/NaCl precipitated pellet by combining resultant 1 mL solution in TRIzol with Lying Matrix B beads (MP Biomedicals) in a 2 mL microcentrifuge tube and lysed using the FastPrep-24 5G (MP Biomedicals) at 6 m/s for 30 seconds (1 cycle). The lysate was centrifuged at 12,000 × g for 1 minute and transferred to a new 2 mL microcentrifuge tube and combined with an equal volume of ethanol (95% - 100%). The resultant mixture was transferred to a Zymo-Spin IC Column (Direct-Zol RNA microprep, Zymo Research) and centrifuged at 12,000 × g for 30 seconds, repeated until entire mixture flowed through column. The column was washed with 400 µL RNA Pre-Wash Buffer. Next, 700 µL RNA Wash Buffer was added to the column and centrifuged for 1 minute and then transferred to a new 1.5 mL microcentrifuge tube. RNA was eluted by adding 20 µL of nuclease-free water into the column and centrifuging for 1 minute.

Viral RNA was quantified using LUNA SARS-CoV-2 RT-qPCR multiplex Assay (New England Biolabs) in a 96-well MicroAmp Reaction Plate with the QuantStudio 5 thermocycler (Thermo Fisher, Carlsbad, CA.). Primer/probe mix (N1/N2/RP) (1 µL), Luna One-step RT-qPCR 4x Mix with UDG (2.5 µL), and nuclease-free water (2.5 µL) were placed into 96-well plate and combined with extracted RNA (4 µL). The plate was sealed and centrifuged at 1,000 g for 30 seconds to push reactions to the bottom of wells. The RT-qPCR detection was programmed for one cycle of (25°C 30 sec, 55°C 10 min, and 95°C 1 min) and 45 cycles of (95°C 10 sec and 60°C 30 sec with plate read), using fluorescence HEX for N1, FAM for N2, and Cy5 for RP targets. A standard curve was generated using diluted synthetic SARS-CoV-2 (Twist Bioscience) at 0, 10, 100, 1000, and 10000 copies. Prior to 15 February 2021, quantification of viral copies obtained from wastewater was not achieved, and thus only presence/absence data were available prior to this date.

### Student COVID numbers and Vaccine information

The number of confirmed cases of COVID-19 among students was obtained from student housing databases and only included students residing on campus. The numbers of student COVID-19 cases for each dorm during each semester were tallied. Total student housing numbers established at the beginning of the semester were used with the number of positive cases to determine incidence rates per semester. Student vaccine data were obtained from ECU COVID-19 vaccine dashboard on the ECU website. The ECU dashboard was a visual representation published on the university’s website used to convey COVID-19 information. The dashboard was used as a tool to help monitor COVID-19 trends across campus.

### Statistical Analysis

Student COVID-19 cases were analyzed for each dormitory on a semester basis to determine the dorms that had the highest number of COVID-19 cases and dorms that had the lowest number of COVID-19 cases. The mean SARS-CoV-2 concentration (virus copies per mL) in wastewater for each dormitory was calculated during the Spring (Feb – May 2021) and Fall (Aug – Nov 2021) semesters. Mean concentrations of SARS-CoV-2 in wastewater from dormitories with relatively high student COVID-19 cases were compared to the concentrations of those with relatively low number of student cases to determine if the differences were statistically significant (p < 0.05). As dormitory capacity differed between Spring and Fall semesters, only single occupancy was allowed during the Spring, “low” and “high” numbers of confirmed COVID-19 student cases differed between the two semesters. Additionally, as classes were predominantly offered online during the Spring, fewer students opted to stay on campus. Thus, relatively high student cases were classified as dorms having greater than 5 cases in the Spring and greater than 10 cases for the Fall (dormitories returned to double occupancy during Fall semester). The normality and linearity of data were evaluated to determine which statistical tests were most appropriate. A chi-square test of independence was performed to determine if there was a statistically significant difference between mean viral copies per mL of sample between dorms with “low” and “high” numbers of confirmed COVID-19 student cases. For nonparametric data, Kruskal-Wallis tests were used to determine significance between data consisting of more than 2 categorical grouping variables, while Mann-Whitney testing were used to analyze significance between 2 samples. Statistical analyses were performed using SPSS (SPSS Institute, Chicago, Ill). SPSS analysis also included a post-hoc pairwise Mann-Whitney with Bonferroni adjustments. Weekly averages for SARS-CoV-2 in wastewater were determined during each semester and any increases in the week-to-week viral concentrations were noted. If a new positive COVID-19 case in a dorm followed an increase in the weekly mean concentration of SARS-CoV-2 in wastewater from that dorm, then the wastewater tests and analyses were considered an accurate predictor of new cases. Student COVID-19 cases documented the week following a high viral concentration were tabulated and used to calculate the percentage of time new cases were identified following high virus counts. Additionally, the relationship between the number of virus copies in wastewater during the spring semester and the number of student cases was evaluated using Pearson’s correlation.

## Results

### Comparison of COVID Cases across dormitories

Overall, there was a total of 367 ECU confirmed cases of COVID-19 within the dormitories during the Spring and Fall semesters of 2021 (Figure 2). The highest number of cases and highest incidence rates were observed during March (Spring) and September (Fall) (Table 1, Table 2). Both of the incidence rises align with campus events that may have precipitated or affected the perceived increase in cases. For example, Spring increases occurred around the time of traditional spring break events for students, while Fall increases occurred during Labor Day weekend, which also coincided with the first home football game. During the Spring semester, the Greene, Scott and Jones dorms contained the highest number of COVID cases (combined total cases, n= 33). The highest mean (M) number of cases for those dorms was M = 11. Subsequently, White, Clement, and Ballard West dorms had the lowest number of COVID-19 cases (n = 9) and the lowest average number of cases per dorm (M = 3). For the Fall semester, sampling results indicated that Clement, Legacy and Fletcher dorms had the highest number of COVID cases (n = 113), and the three dorms averaged 37.7 cases each. Garrett, Fleming and Jarvis had the lowest number of cases (n = 21) and averaged 7 cases per dorm.

**Figure 2.**
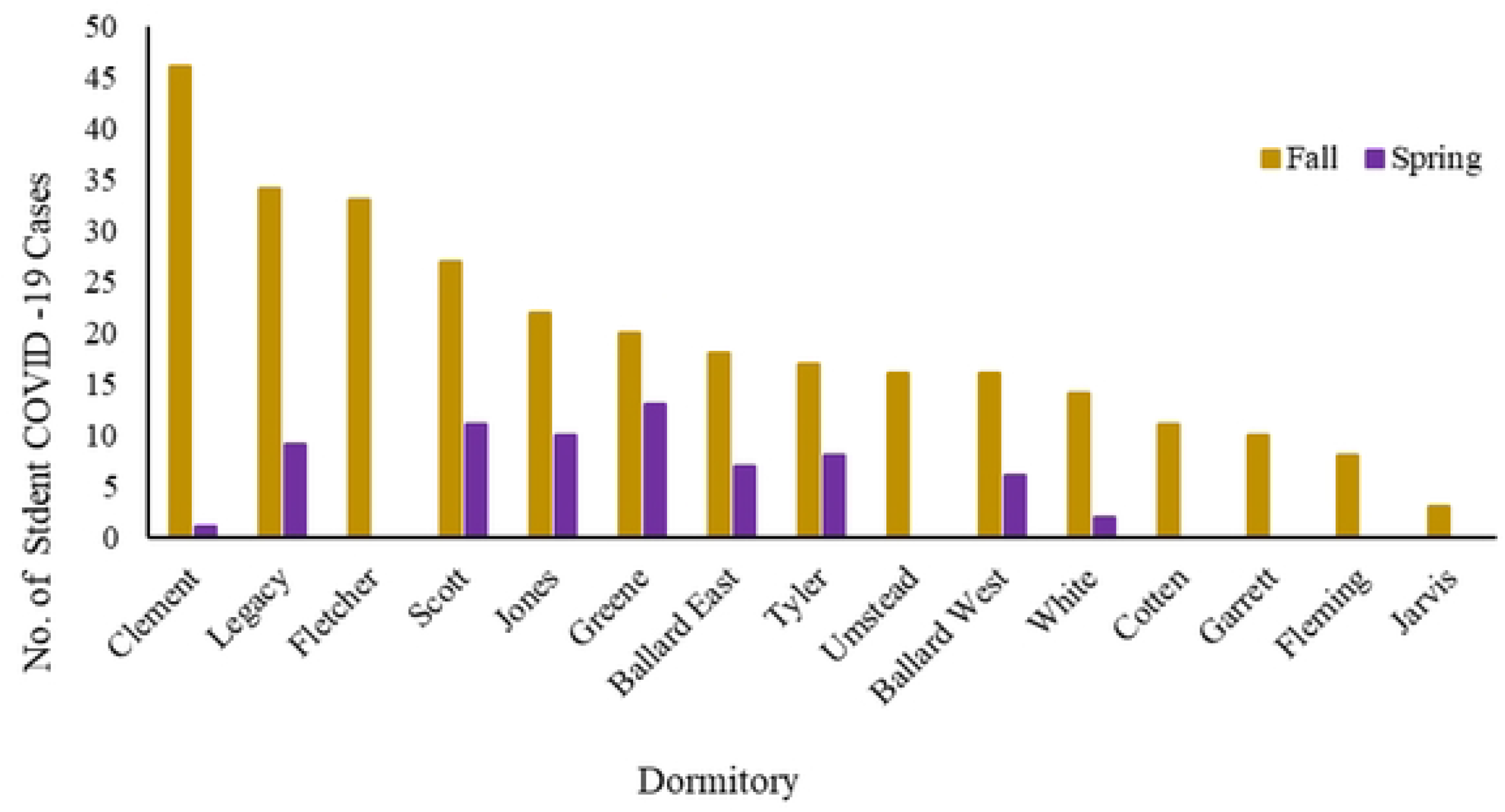
Total number of COVID-19 cases among students residing in dorms during the Spring and Fall 2021 Semesters.

**Table 1.**
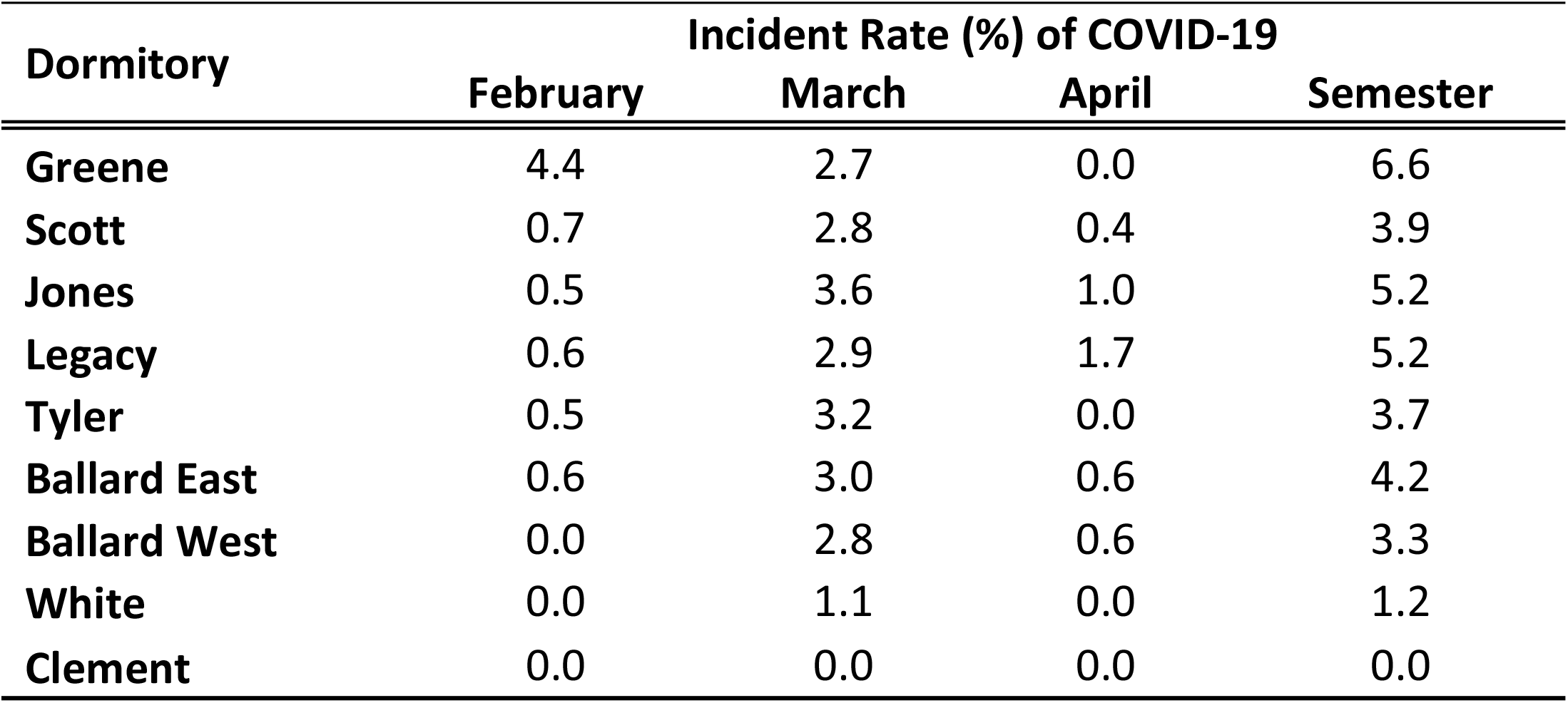
COVID Incidence Rates for Sampled Dormitories Between 15 February 2021 and 1 May.

**Table 2.**
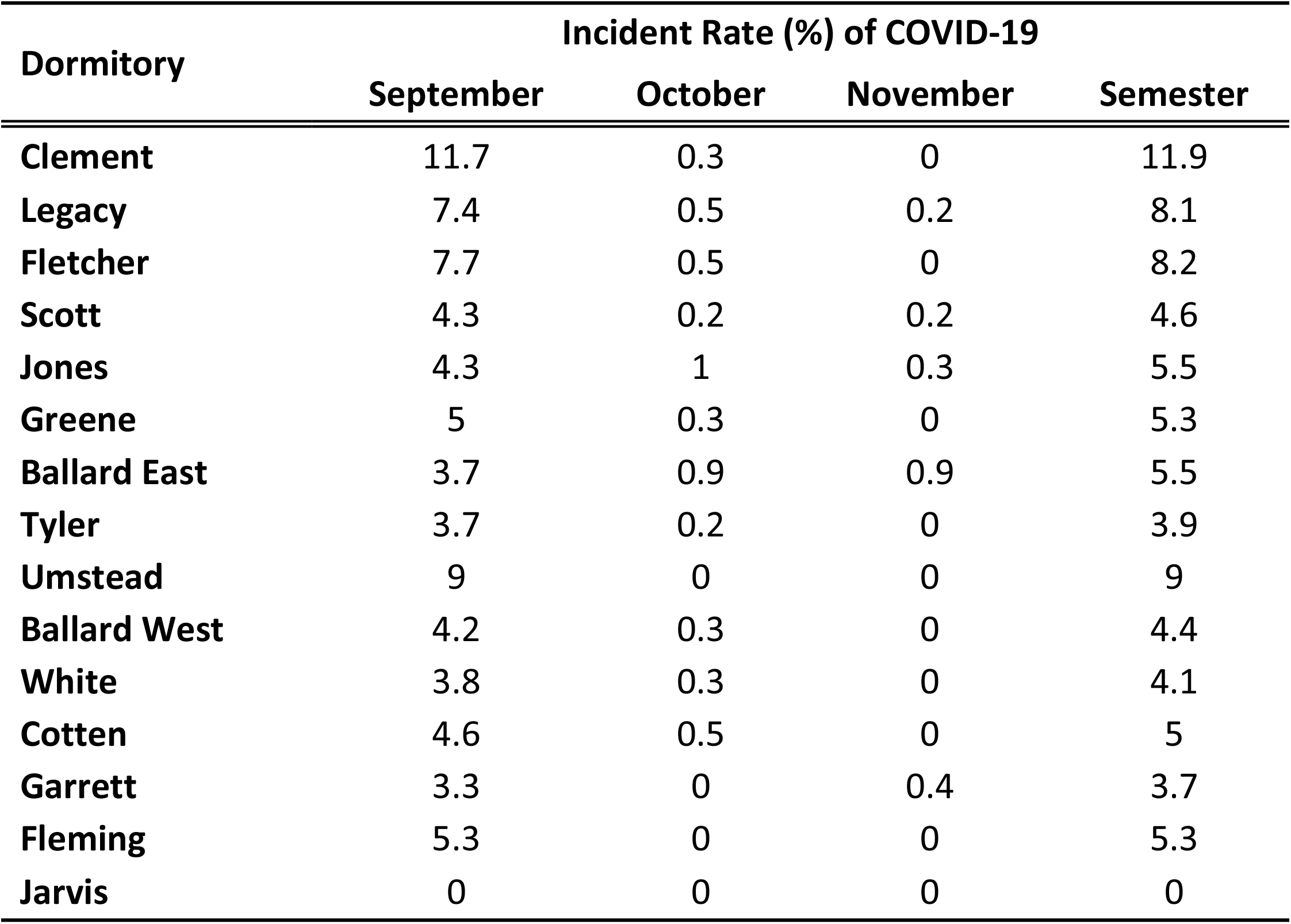
COVID incidence rates for sampled dormitories between 30 August 2021 and 19 November 2021.

Thus, there were large differences in the number of COVID-19 cases between dorms, with some having 3 or more times as many as others. However, not all dorms housed the same number of students. Additionally, there were differences in maximum student housing occupancy for the semesters. For example, during the Spring 2021 semester student housing was limited to one person residency per dorm room, which was approximately half capacity. The following semester (Fall 2021) those restrictions were lifted, and rooms could support normal occupancy, thus allowing multiple students to share a room. It may be possible that the number of students in the dorms affected the number of COVID cases captured.

We also sought to determine whether the number of students in dorms could be used as a predictor of COVID-19 cases. Using linear regression, results showed that while the model generated was significant, (p = 0.006), it may only explain 10.2% of the variance. Overall, the number of students in dorms did significantly predict the number of COVID-19 cases detected (ß_1_ = -0.026, p = 0.006), with a final predictive model = -1.24 + (0.023*Number of Students in Dorm).

### SARS-CoV-2 Concentration in Wastewater

Concentrations of SARS-CoV-2 in wastewater exhibited temporal variation between semesters. Overall, changes in virus gene copies throughout both semesters were observed. Large increases in SARS-CoV-2 concentrations in wastewater during the early Spring semester were noticed. The City of Greenville was also conducting wastewater-based epidemiology during the same time frame. While the magnitude of wastewater flow and viral concentrations were different between the City of Greenville and ECU, both reported large increases in SARS-CoV-2 concentrations in wastewater after the start of the Spring and Fall semesters during February and September, respectively (Figure 3A, Figure 3B). The September increases corresponded with a fully re-opened ECU campus and related activities (sports games, Labor Day festivities, etc.,).^14^ Near the start of the Spring semester in February 2021, concentrations of SARS-CoV-2 in wastewater increased between 20 – 39% for the City of Greenville while ECU experienced an increase of 11.2%. Similarly, near the start of the Fall semester of 2021, ECU experienced a 96.3% increase in SARS-CoV-2 concentrations in dorm wastewater while the City of Greenville experienced a 70 – 89% increase.

**Figure 2A.**
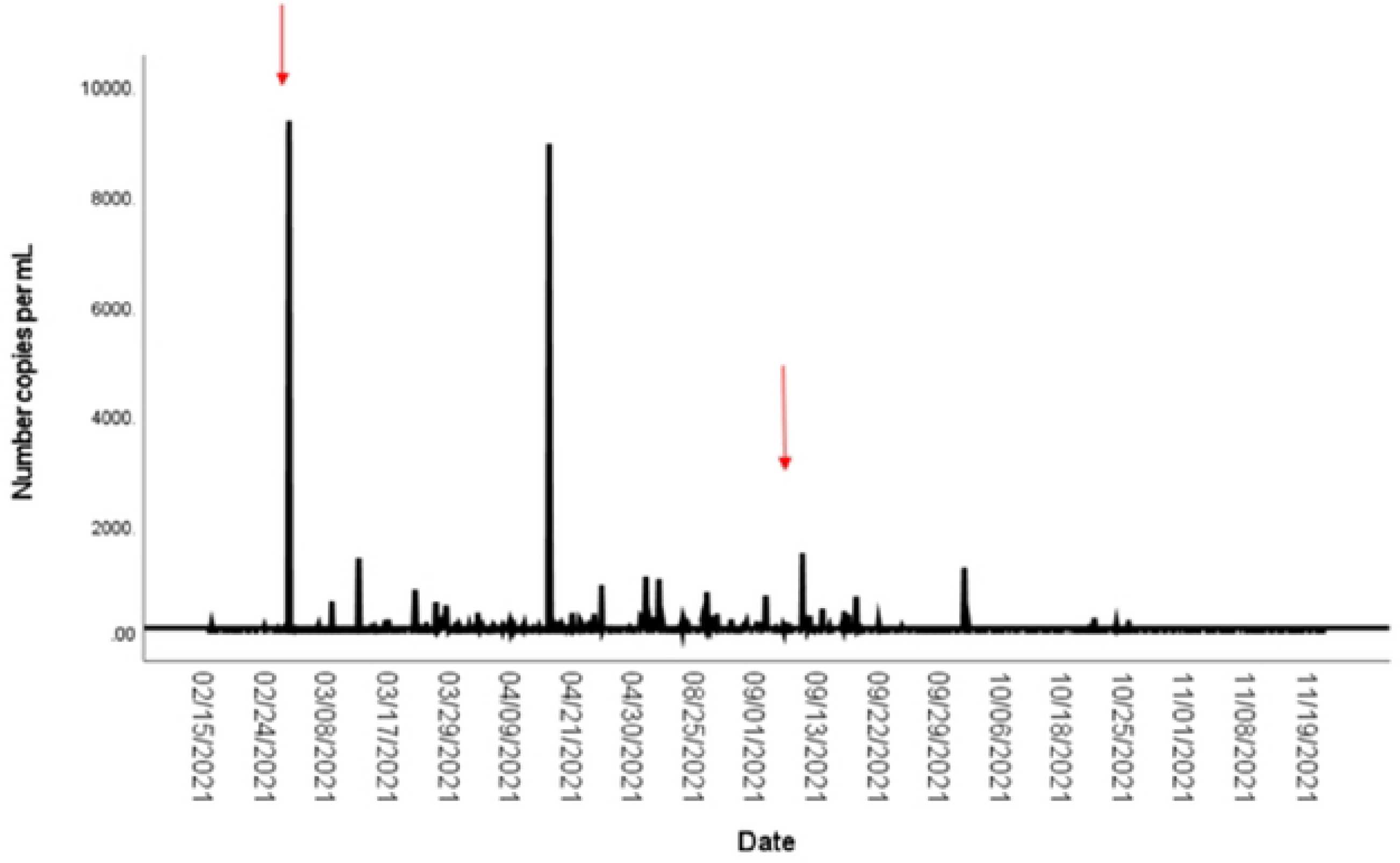
SARS-CoV-2 in dormitory wastewater samples Collected throughout the Spring 2021 and Fall 2021 Semesters. Red arrows denote times when the City of Greenville also saw noticeable increases in viral copies of COVID-19 in wastewater.

**Figure 3B.**
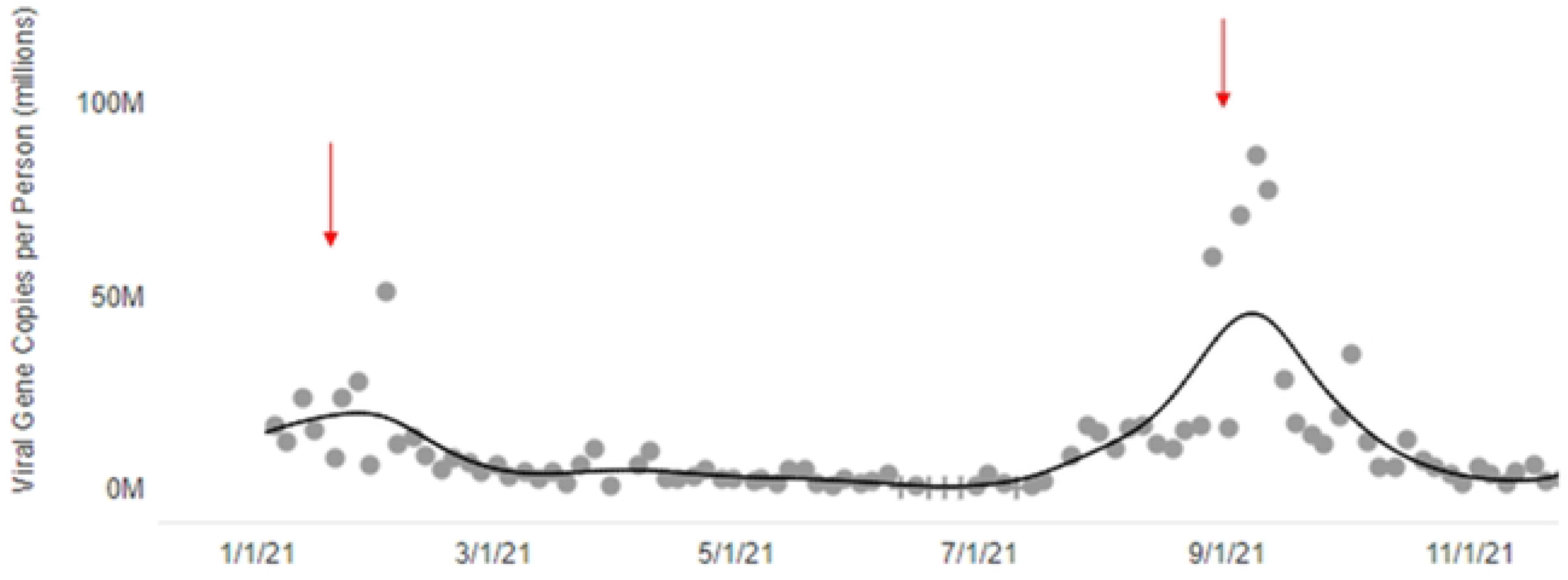
SARS-CoV-2 in wastewater samples for the City of Greenville during the 2021 Year. Red arrows denote times when the East Carolina University also saw noticeable increases in COVID-19 virus in wastewater. Modified from NC DHHS (2022) wastewater sampling dashboard.

### Comparison of SARS-CoV-2 Concentrations in Wastewater from Dormitories with Higher and Lower Student Cases of COVID-19

During the Spring 2021 semester, student COVID-19 testing showed the highest number of confirmed cases were in the dormitories of Scott (n = 8) and Jones (n = 9). White (n = 2) and Clement (n = 1) had the lowest number of positive COVID-19 cases. During the Spring semester, the mean concentration of SARS-CoV-2 among dormitories with the highest number of student cases was 111.1 viral copies per mL, while dormitories with the lowest number of cases contained a mean concentration of 7.3 viral copies per mL. This difference was statistically significant (U= 10.0, *p*= 0.002). The results showed a statistically significant positive correlation between the two, (r = 0.642, N= 20, *p*= 0.002). This indicates that when the number of virus copies increased there was a corresponding increase in the number of student cases during the Spring.

During the Fall 2021 semester, the dorms with the highest number of students with COVID-19 were Clement (45) and Legacy (31). Conversely, the dorms with the lowest numbers of COVID-19 cases were Fleming (8) and Jarvis (3). The mean concentration of SARS-CoV-2 for the dormitories with the highest number of COVID-19 cases was 49.3 viral copies per mL, while the dorms with the lowest number of COVID-19 cases had a mean of 3.6 viral copies per mL. This difference was statistically significant (U= 34; *p*= 0.023). A Pearson correlation performed on the Fall semester data also showed a significant positive relationship between virus copies and cases, (r = 0.522, N= 34, p= 0.013).

### Wastewater Sampling and Determination of New Cases

Overall, of the 830 samples processed and analyzed, 594 (71%) tested positive for the virus. Prior to 15 February 2021, quantification of viral copies obtained from wastewater was not achieved, and thus only presence/absence data were available. However, in the week leading up to February 15, 2021, positive samples for 77% of dorms were reported. Student COVID-19 cases were confirmed following this sampling period with 55% of dorms reporting cases and some dorms having multiple cases. Typically, ECU celebrates spring break around the 1^st^ week of March (corresponding here 1 March – 7 March). During 2021, the University did not hold a traditional spring break, but there was a decrease in the number of students with COVID-19 during that week. It may be possible that some students in keeping with past traditions left campus (as classes were virtual) and returned later. Additionally, in lieu of a full “Spring Break” week for students, the university sponsored a “Spring Festival” on 10 March. A 7-fold rise in cases after the festival and extending through the end of March 2021 was observed (Figure 4)

**Figure 4.**
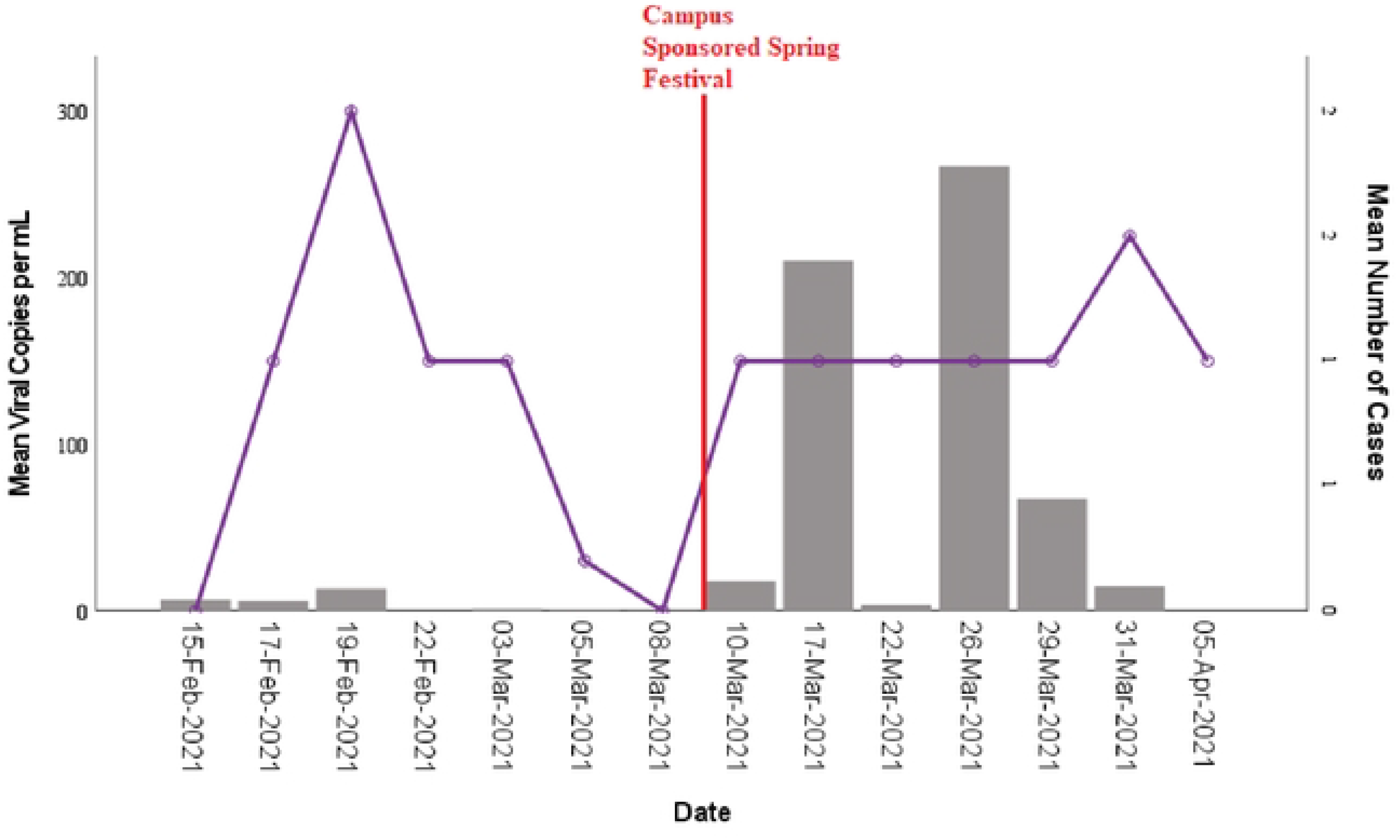
Time-lapse of mean SARS-CoV-2 virus copy per mL extracted from wastewater and students COVID-19 cases in dormitories during the spring semester. Bars represent mean virus copy per mL and lines represent student cases.

An increase in virus concentrations in wastewater at the beginning of the Fall semester corresponded with an increase in the number of student cases (Figure 5). More specifically, at the beginning of semester, testing showed a rise in viral copies of SARS-CoV-2 in wastewater starting on 23 August 2021 (208.2 copies per mL) to 3 September 2021 (1070 copies per mL) which coincided with a 97.5% increase in the number of students testing positive for COVID-19. The number of positive students peaked on 4 September 2021. Students that tested positive for COVID-19 were removed from their respective dorms and placed into isolation dorms. During that time in isolation dorms, concentrations of viral copies in wastewater from the home dorms of the students declined 67.2%. Virus copies may have also fallen during this time due to students traveling for the Labor Day holiday on 6 September 2021. However, when students began returning to their original dorms from isolation (∼10 days, around 13 to 17 September 2021), a 74.1% rise in virus concentration in wastewater sampled from their original dorms was observed, possibly because those students were still shedding the virus. This may explain the persistence of relatively high concentrations of SARS-CoV-2 in wastewater even as the number of student cases declined.

**Figure 5.**
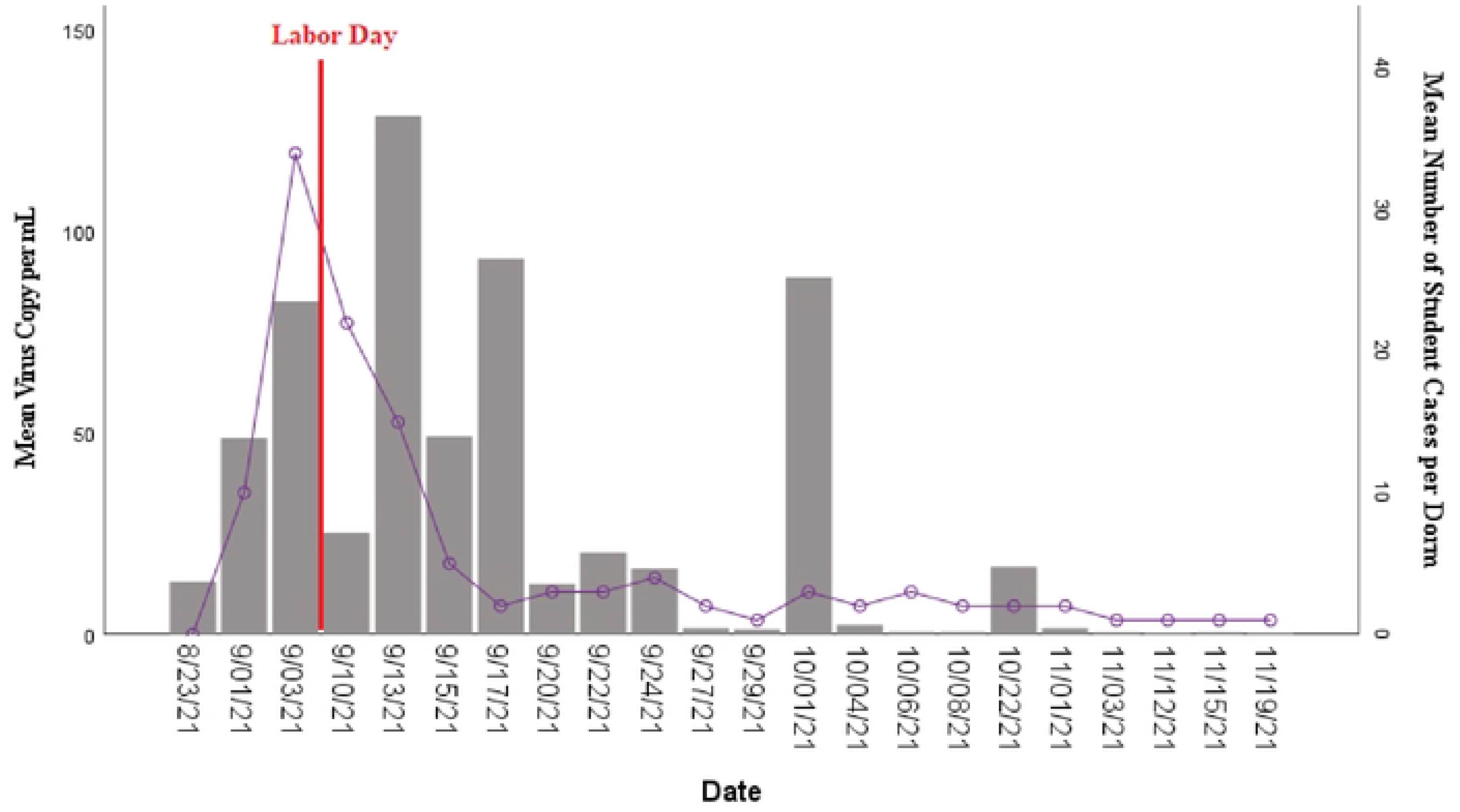
Time-lapse of mean SARS-CoV-2 virus copy per mL extracted from wastewater and students COVID-19 cases in dormitories during the fall semester. Bars represent mean virus copy per mL and lines represent student cases.

Studies have shown persons may shed the virus many days prior to symptom onset (typically persons seek testing after experiencing symptoms). ^15, 16^ For this reason, new COVID cases (n= 1) in students that occurred within a week following a high viral count in wastewater were recorded and the frequency of positive cases calculated. Overall, about 60% of the time at least 1 new case of COVID-19 was observed following a SARS-CoV-2 concentration of 9.2 copies per mL in wastewater sampled from a dorm. There was great variability between the Spring and Fall semesters regarding the results though. During the Spring semester, when wastewater samples from dorms yielded at least 9.2 SARS-CoV-2 viral copies per ml there was at least 1 new COVID-19 case in dorms within the following week during 47% of the times tested. New cases of students with COVID-19 were observed the subsequent week following a testing that yielded 9.2 copies of the virus 73.8% of time during the Fall semester. Additionally, as some students recognized they may have had an increased chance of exposure, they may have voluntarily removed themselves from dormitories which may have also impacted the results from that time period, with new cases not showing as students left before being tested. When the data are reviewed based on percentage increases in SARS-CoV-2 concentrations in wastewater and identification of new cases, less variability is observed between semesters. More specifically, during the Spring semester, when the concentration of SARS-CoV-2 in wastewater increased by 24.4% in a dormitory, a new case of COVID-19 was observed 68.8% of the time the week following the increase. During the Fall semester, a new case of COVID-19 was observed 64.3% of time the week following a 17.6% increase in SARS-CoV-2 concentration in wastewater.

### Saliva Surveillance

Students, faculty, and staff participated in saliva surveillance particularly if they had been exposed to someone who had COVID-19. During the Spring semester, it was required that all student athletes and 25% of students who lived on-campus participate in saliva testing. As the university fully opened in Fall 2021, students enrolled in face-to-face classes, residing on campus and participating in NCAA athletics were required to be a part of the routine surveillance. At the beginning of the Fall semester at least 50% of on campus students were required to be tested weekly. However, as the number of cases increased, it was determined beginning 1 September 2021, that surveillance testing frequency would also increase with unvaccinated students residing in residence halls. Unvaccinated students living in dorms had to undergo weekly testing, with some testing being PCR testing. Saliva surveillance of students living in residence halls during 1 September 2021 through 30 September 2021 were analyzed. During this time, the University processed 2820 samples with 16 samples (0.56%) testing positive. During the first 2 weeks the trend in number of positive saliva tests and the mean concentrations of SARS-CoV-2 in wastewater was similar. An initial drop in both the mean virus concentrations in wastewater and the number of positive saliva tests over the first week (3 September to 11 September 2021) were followed by at least 2-fold increases in virus concentrations in wastewater and positive saliva tests the next week (Figure 6). However, the trends for mean virus concentrations in wastewater and number of positive saliva tests diverged in the last week of September 2021. A Spearman’s rho correlation determined an overall weak association between SARS-CoV-2 obtained from wastewater and positive saliva samples (*r*= 0.200, *p*= 0.800). The overall weak association between virus concentrations in wastewater and the number of positive saliva samples may be because the sample populations were different. All students that had face to face classes including those that lived off campus were included in the saliva testing, while the wastewater analyses were only applicable for students that lived on campus. Thus, the sample sizes and populations were different.

**Figure 6.**
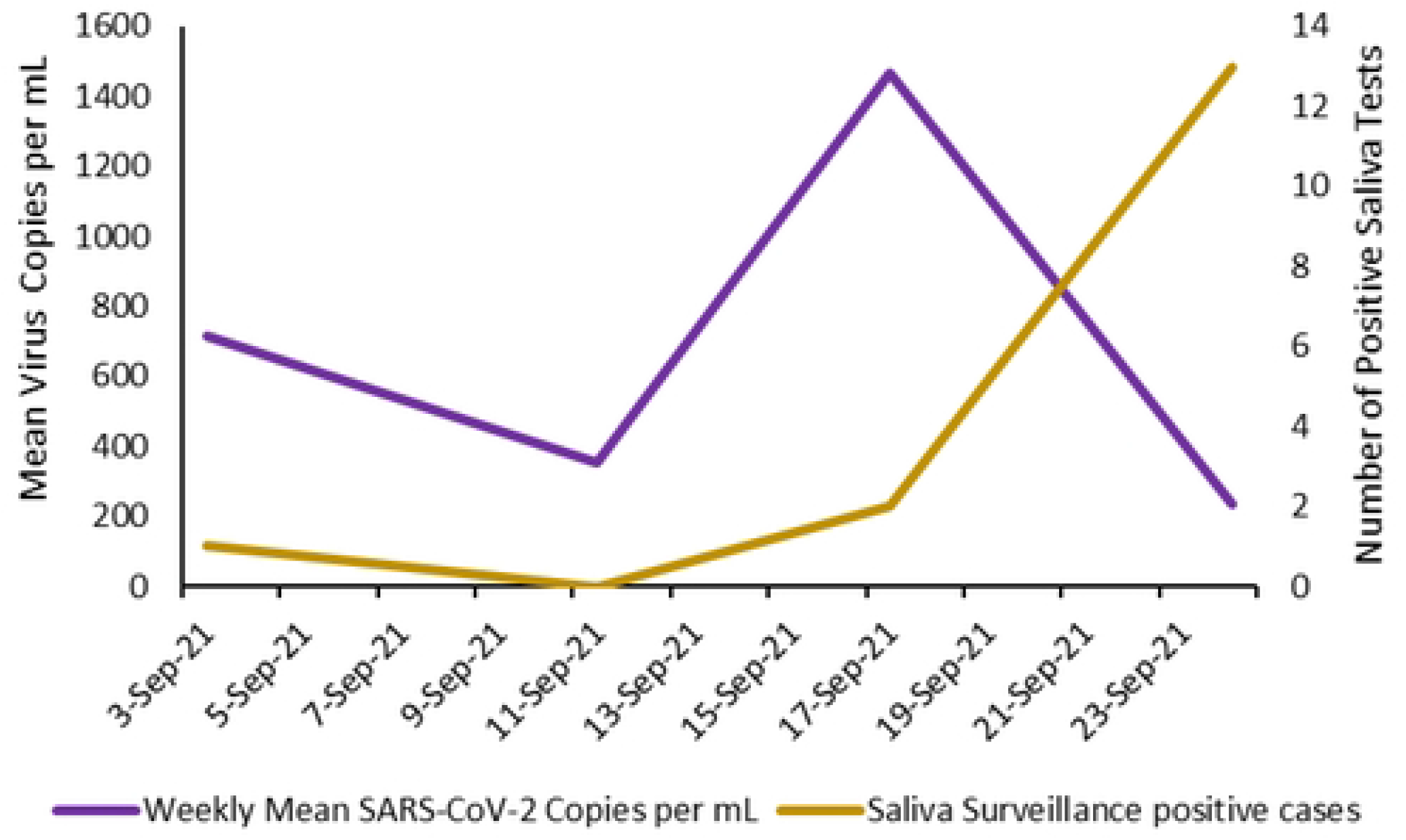
Positive Saliva Surveillance Samples compared to mean SARS-CoV-2 copies per mL obtained from wastewater.

## Discussion

The COVID-19 pandemic was unprecedented in that it led governmental officials to shut down and/or pivot routine services. This included educational institutions and many private businesses. Colleges and universities were tasked with developing and/or incorporating new ways to monitor SARS-CoV-2 on their campuses to keep students, faculty, and staff safe as services and operations were gradually restored. WBE was a tool used by ECU to help prevent the spread of COVID-19. Wastewater solids tend to be negatively charged and SARS-CoV-2 is a positive-sense single stranded virus.^17.18^ Thus, SARS-CoV-2 may sorb to wastewater solids and the wastewater may be used as an indicator of the virus. This study aimed to determine if wastewater-based epidemiology was effective as an environmental public health surveillance system to help control COVID-19 outbreaks on college campuses.

An objective of this study was to determine if wastewater monitoring could be used to reduce the likelihood of outbreaks by triggering student testing and isolation when SARS-CoV-2 concentration in wastewater spiked and subsequent testing for related dorms showed positive cases. The CDC has stated that COVID-19 outbreaks may be foreshadowed by results of wastewater tested 7 days earlier.^19^ For the current study, it was noted that at specific times during each semester when a rise in virus concentrations in wastewater was observed an increase in the number of COVID-19 cases in on-campus students typically (60% of times) followed 5-7 days later. This same trend was noted when observing percentage increases in virus concentration in wastewater, where overall new COVID cases were found ∼ 71.2% of times. Thus, WBE may help reduce the likelihood of future outbreaks as this potential 7-day period may provide meaningful time for public health officials in contacting/identifying potential cases and placing those cases in isolation. This would be of tremendous aid in helping to mitigate the spread of disease, as this would possibly help to reduce the number of potential contacts for the infected persons.

However, it is important to note that changes in guidelines and protocols may affect how data are used to predict cases. For example, during the Fall semester, students were allowed to have visitors that did not live in the dormitories. This was not allowed during the Spring semester. It is possible that visitors with COVID-19 contributed to viral concentrations in wastewater to the tested dorms. This may cause a large enough rise in viral concentration to trigger mass PCR/swab testing without yielding new cases from that specific dorm, thereby negatively affecting the accuracy of WBE in predicting future cases. Researchers and decisionmakers need to also be aware of those returning from quarantine and isolation. As guidelines and timelines change, more people could be introduced back into the dorms potentially shedding the virus. Studies have shown that the medium detectable timeframe for the virus in stools was 17 (11-32) days, but some patients may shed up to 59 days.^20, 21^ Students at ECU were allowed to leave quarantine and return to their dorms after 10 days. It is not practical for infected students to isolate for 32 - 59 days due to logistical and social concerns. The isolation dorm does not have the capacity to house a substantial percentage of the student population for 1 – 2 months. Furthermore, long-term isolation could excessively burden university staff and administration due to the time and resource investment required to regulate compliance and resolve conflicts with students and their families. Therefore, those returning students may have continued shedding detectable SARS-CoV-2 in wastewater, increasing the virus concentrations, and triggering “false alarms” regarding the need for saliva testing.

Persons may also shed the virus at different rates. Studies have shown persons affected with coronaviruses typically have lower rates of viral shedding in the initial days, with peak shedding 12-14 days after disease onset.^22^ However, viral shedding is known to occur prior to the onset of symptoms for SARS-CoV-2.^20, 23^ These issues may impact an important component of using wastewater surveillance as a predictor tool. Public health officials have to be able to set viral thresholds that may trigger additional testing. Skewing of this data may affect the ability of public health professionals to use one standard isolation protocol. Thresholds for triggering actions such as swarm testing may need to be set for individual entities and may need to be fluent and allow for change during different time periods of an outbreak.

An additional challenge may be testing compliance. The timing and ability to test suspected persons is important in mitigating spread of the virus. Students often have differing schedules, and this may impede swarm testing. Some may also feel testing is not important or they have an aversion to how the test is performed. Politicizing of COVID-19 virus and vaccinations resulted in polarization of the issue, which may have added to unease of virus discussion among healthcare professionals. For these reasons, it is imperative that colleges and universities develop plans to help increase the percentage of students who are compliant with testing protocols.

Despite these challenges, wastewater surveillance may be an effective tool as part of a comprehensive surveillance system for use by colleges, universities, and other institutions. Persons who are asymptomatic or have mild symptoms may not realize they are carriers and/or are spreading the virus. Asymptomatic patients may forego contact tracing, which may lead to underestimates of the case numbers and may exacerbate the viral spread. Thus, detection using WBE may be helpful in identifying these cases. Wastewater-based surveillance may also be useful to determine fluctuations in outbreaks once they become endemic in an area. As viruses mutate, the virulence and transmissibility may be affected.^24^ Upticks in cases may alert officials to changes in the virus.

Additionally, use of wastewater viral data may help inform officials of the efficacy of existing mitigation strategies and protocols and whether current strategies need to be adjusted. It may also be used to implement focused surveys or questionnaires to determine compliance with existing strategies and whether compliance is an issue. Using wastewater-based epidemiology as an early warning system may allow public health officials to evaluate the spread or potential future spread of a disease.

### Limitations

While this study aimed to analyze wastewater-based epidemiology in public health surveillance, several limitations were encountered. Current research has shown SARS-Cov-2 may be extracted from wastewater; however, data are lacking on the concentration of the virus that is typically shed in feces (i.e., illness duration and differences in persons). Thus, it was possible for us to quantify SARS-CoV-2 concentrations in wastewater over time, however we were not able to estimate or determine the exact number of virus copies that would correspond to an infected individual. Researchers have also noted as time passed during the semester, it was difficult to determine SARS-CoV-2 virus copies shed from persons returning from quarantine and new cases in the same dorm. Additionally, issues such as clogged autosamplers resulting from inappropriate student waste disposal may have affected the ability to collect full-volume samples. Student compliance with regards to saliva testing also inhibited timely testing at times during the semester, affecting case identification. We also were not able to ascertain flow rate from dormitories into the sewer system, which could be used to determine flow-weighted concentrations. These data can be valuable in assessing wastewater concentrations since certain activities (e.g., washing clothes, dishwashing, showering) may generate large flow volumes without contributing as much virus as toilets or sinks. Thus, sudden declines in SARS-CoV-2 concentrations that are not explainable via public health data could be attributed to differences in flow on sampling day.

## Data Availability

All relevant data are within the manuscript and its Supporting Information files.

## Acknowledgments

We would like to acknowledge ECU facility services, ECU Student Affairs, ECU Student Health, Cavanagh Lab, Fallon Lab, and the Wastewater lab for their immeasurable help on this project.

## References

1. (CDC) Centers for Disease Control and Prevention. Public Health Training: Introduction to Public Health Surveillance. 2018. Accessed at: https://www.cdc.gov/training/publichealth101/surveillance.html

2. Groseclose SL, Buckeridge DL. Public health surveillance systems: Recent advances in their use and evaluation. 2017. Annual Review of Public Health. 38: 57–79

3. Nsubuga P, White ME, Thacker SB, Anderson MA, Blout SB, Broome CV, Chiller TM, Espitia V, Imtiaz R, Sosin D, Stroup DF, Tauxe RV, Vijayaraghavan M, Trostle M. Public Health Surveillance: A Tool for Targeting and Monitoring Interventions. Disease Control Priorities in Developing Countries. 2nd edition. Washington (DC). Oxford University Press, New York. 2006. Accessed at. https://www.ncbi.nlm.nih.gov/books/NBK11770/

4. Balajee AS, Salyer SJ, Greene-Cramer B, Sadek M, Mounts AW. The practice of event-based surveillance: concepts and methods. Global Security: Health, Science and Policy. 2020. 6(1): 1–9

5. Devault DA, Karolak S. Wastewater-based epidemiology approach to assess population exposure to pesticides: a review of a pesticide pharmacokinetic dataset. Environmental Science and Pollution Research. 2020. 27: 4695–4702

6. Zuccato E, Chiabrando C, Castiglioni S, Calamari D, Bagnati R, Schiarea S, Fanelli R. Cocaine in surface waters: a new evidence-based tool to monitor community drug use. Environmental Health. 2005.4: 14. doi :10.1186/1476-069X-4-14

7. Kankaanpää A, Ariniemi K, Heininen M, Kuoppasalmi K, Gunnar T. Current trends in Finnish drug abuse: Wastewater based epidemiology combined with other national indicators. Science of the Total Environment. 2016. doi: 10.1016/j.scitotenv.2016.06.06

8. Yang Z, Kasprzk-Hordern B, Frost CG, Estrela P, Thomas KV. Community sewage sensors for monitoring public health. Environmental Science and Technology. 2015. 49(10): 5845–5846

9. Sims N, Kaspryzk-Hordern B. Future perspectives of wastewater-based epidemiology: Monitoring infectious disease spread and resistance to the community level. Environmental International. 2020. doi: 10.1016/j.envint.2020.105689

10. Wang MY, Zhao R., Gao L J, Gao X F, Wang DP, Cao JM. SARS-CoV-2: Structure, Biology, and Structure-Based Therapeutics Development. Frontiers in cellular and infection microbiology. Frontiers in Cellular and Infection Microbiology. 2020. 10: 587269. https://doi.org/10.3389/fcimb.2020.587269

11. Betancourt WQ, Schmitz BW, Innes GK, Prasek SM, Pogreba Brown KM, Stark ER, Foster AR, Sprissler RS, Harris DT, Sherchan SP, Gerba CP, Pepper IL. COVID-19 containment on a college campus via wastewater-based epidemiology, targeted clinical testing and an intervention. Science of the Total Environment 2021. 779: doi:10.1016/j.scitotenv.2021.146408

12. Crowley M. a. 2020. Accessed at: https://www.norwich.edu/news/2858-norwich-university-wastewater-based-epidemiology-inititative

13. Gibas C, Lambirth K, Mittal N, Islam MA, Barua VB, Brazell LR, Hinton K, Lontai J, Stark N, Young I, Quach C, Russ M, Kauer J, Nicolosi B, Chen D, Akella S, Tang W, Schlueter J, Munir M. Implementing building-level SARS-CoV-2 wastewater surveillance on a university campus. Science of the Total Environment. 2021. 782: doi: 10.1016/j.scitotenv.2012146749

14. (NCDHHS) North Carolina Department of Health and Human Services. NCDHHS Shares Updated Rollout Plan for COVID-19 Vaccinations. 2020. Accessed at: https://www.ncdhhs.gov/news/press-releases/2020/12/30/ncdhhs-shares-updated-rollout-plan-covid-19-vaccinations

15. He X, Lau EHY, Wu P, Deng X, Wang J, Hao X, Chung YC, Wong JY, Guan Y, Tan X, Mo X, Chen Y, Liao B, Chen W, Hu F, Zhang Q, Zhong M, Wu Y, Zhao L, Zhang F, Cowling BJ, Li F, Leung GM. Temporal dynamics in viral shedding and transmissibility of COVID-19. Nature Medicine. 2020. (26): 672–675

16. Lewis NM, Duca LM, Marcenac P, Dietrich EA, Gregory CJ, Fields VL, Banks MM, Rispen JR, Hall A, Harcourt JL, Tamin A, Willardson S, Kiphibane T, Christensen K, Dunn AC, Tate JE, Nabity S, Matanock AM, Kirking HL. Characteristics and Timing of Initial Virus Shedding in Severe Acute Respiratory Syndrome Coronavirus 2, Utah, USA. Emerging Infectious Disease. 2021.27(2): 352-359

17. Ye Y, Ellenburg RM, Graham KE, Wigginton KR. Survivability, Partitioning, and recovery of Enveloped Viruses in Untreated Municipal Wastewater. Environmental Science Technology. 2016. 50(10): 5077–5085

18. Jackson CB, Farzan M, Chen B, Choe H. Mechanisms of SARS-CoV-2 entry into cells. National Review of Molecular Cell Biology. 2022. 23(1): 3–20

19. Crowley M. Northfield, Norwich University partner, invited to join national wastewater surveillance system. Norwich University Newsroom. 2022. Accessed at: https://www.norwich.edu/news/3602-norwich-university-northfield-wastewater-based-epidemiology-initiative

20. Jones DL, Baluja MQ, Graham DW, Corbishley A, McDonald JE, Malham SK, Hillary LS, Conner TR, Gaze WH, Moura IB, Wilcoz MH, Farkas K. Shedding of SARS-CoV-2 in feces and urine and its potential role in person-to-person transmission and the environment-based spread of COVID-19. Science of the Total Environment. 2020. 749: doi:10.1016/j.scitotenv.2020.141364

21. Huang J, Mao T, Li S, Wu L, Xu X, Li H, Xu C, Su F, Dai J, Shi J, Cai J, Huang C, Lin X, Chen D, Lin X, Sun B, Tang S. Long Period Dynamics of Viral Load and Antibodies for SARS-CoV-2 Infection: An Observational Cohort Study. 2020.Accessed at: medRxiv.doi: https://doi.org/10.1101/2020.04.22.20071258

22. Cheng P.K.C., Wong D.A., Tong L.K.L., Ip S.M., Lo A.C.T., Lau C.S., Yeung E.Y.H., Lim W.W.L. Viral shedding patterns of coronavirus in patients with probable severe acute respiratory syndrome. Lancet. 2004;363:1699–1700. doi: 10.1016/S0140-6736(04)16255-7

23. Wei WE, Li Z, Chiew CJ, Yong SE, Toh MP, Lee VJ. Presymptomatic transmission of SARS-CoV-2-Singapore. Morbidity and Mortality Weekly Report. 2020. 69:411–415

24. Shao W, Li X, Goraya MU, Wang S, Chen J-L. Evolution of Influenza A Virus by Mutation and Reassortment. International Journal of Molecular Science. 2017. 18(8): 1650. DOI: 10.3390/ijms18081650

